# Xenomonitoring of Lymphatic filariasis and risk factors for transmission on the Kenyan coast

**DOI:** 10.1101/2024.01.23.24301642

**Authors:** Brian Bartilol, Lawrence Babu, Karisa Garama, Jonathan Karisa, Alice Kamau, Charles Mwandawiro, Caroline Wanjiku, Charles Mbogo, Marta Maia, Joseph Mwangangi, Martin Rono

## Abstract

Lymphatic filariasis (LF) is an infectious neglected tropical disease caused by a mosquito-borne nematode and is a major cause of disability. In 2022, it was estimated that 51 million people were infected with LF. In Kenya filariasis is endemic along the entire coastal strip. The main vectors are *Anopheles funestus* and *Anopheles gambiae* in rural areas and *Culex quinquefaciatus* mosquitoes in urban areas.

In 2022, mosquitoes were collected from Kilifi, Kwale and Taita-Taveta counties which are located within the LF endemic region in Kenya. Subsequently, genomic Deoxyribonucleic acid (DNA) was then extracted from these mosquitoes for speciation and analysis of *W. bancrofti* infection rates. The impact of socio-demographic and household attributes on infection rates were assessed using generalized estimating equations.

A total of 18,121 mosquitoes belonging to *Culex (*n = 11,414) and *Anopheles* (n = 6,707) genera were collected. Morphological identification revealed that Anopheline mosquito were dominated by *An. funestus* (n = 3,045) and *An. gambiae* (n = 2,873). *Wuchereria bancrofti* infection rates were highest in Kilifi (35.4%; 95% CI 28%-43.3%, n = 57/161) and lowest in Taita Taveta (5.3%; 95% CI 3.3%-8.0%, n = 22/412). The major vectors incriminated are *An. rivulorum, An. funestus* sensu stricto and *An. arabiensis*. The risk of *W. bancrofti* infection was significantly higher in *An. funestus* complex (OR = 18.0; 95% CI 1.80-180; p = 0.014) compared to *An. gambiae* (OR = 1.54; 95% CI 0.16-15.10; p = 0.7). Additionally, higher risk was observed in outdoor resting mosquitoes (OR = 1.72; 95% CI 1.06-2.78; p = 0.027) and in homesteads that owned livestock (OR = 2.05; 95% CI 1.11-3.73; p = 0.021). Bednet (OR = 0.39; 95% CI 0.12-1.32; p = 0.13) and poultry ownership (OR = 0.52; 95% CI 0.30-0.89, p = 0.018) seems to provide protection.

*Anopheles funestus* complex emerged as the primary vectors of lymphatic filariasis along the Kenyan coast. These findings also highlight that a significant portion of disease transmission potentially occurs outdoors. Therefore, control measures targeting outdoor resting mosquitoes such as zooprophylaxis, larval source management and attractive sugar baits may have potential for LF transmission reduction.

**Author summary:** Lymphatic filariasis (LF) in the African continent is mainly caused by a mosquito-borne nematode: *Wuchereria bancrofti*. In urban areas transmission is mainly by *Culex quinquefaciatus* whereas in rural areas it is dominated by *Anopheles funestus* and *Anopheles gambiae* mosquitoes. We investigated the vectorial systems for LF in rural coastal Kenya and factors associated with the risk of diseases transmission in the region. We identified *An. funestus* sensu lato sibling species *An. rivulorum* and *An. funestus* sensu stricto as the dominant vectors of lymphatic filariasis along the Kenyan coast. We also show that a higher proportion of transmission is likely to take place outdoors necessitating the implementation of vector control strategies that target exophilic mosquitoes such as zooprophylaxis and larval source management. Factors associated with transmission of LF include ownership of livestock and houses made of natural materials such as thatched roof and mud walls. Bednet and poulty ownership were associated with protection. We also highlight the importance of molecular xenomonitoring in the surveillance of lymphatic filariasis, because of its’ non-invasive nature and potential for incriminating new vectors of lymphatic filariasis.

## Introduction

Lymphatic filariasis (LF) is an infectious neglected tropical disease caused by a mosquito-borne nematode: *Wuchereria bancrofti*. Globally LF accounts for 51 million cases with approximately 863 million people in 47 countries still at risk. Clinical symptoms of LF are hydrocele, lymphedema and adenolymphangitis . At an advanced stage, lymphedema develops into elephantiasis which is characterized by swollen body parts (mainly legs, genitals, arms and breasts) and disfiguration that results in sociopsychological problems for patients and their families. In sub-Saharan Africa, filariasis is transmitted to humans by mosquitoes of the genus *Anopheles* and *Culex*. In urban areas transmission is mainly by *Culex quinquefaciatus* whereas in rural areas it is dominated by *Anopheles funestus* and *Anopheles gambiae* mosquitoes (1,2). Transmission occurs through bites from female mosquitoes infected with L3 larvae resulting from microfilariae ingested from infected humans. Once they penetrate the skin, the L3 larvae migrate to the lymphatic system where they mature into adult worms causing disruption in normal circulation leading to clinical symptoms previously described. The worms also produce microfilariae that migrate back to the blood stream and get ingested by a mosquito during a subsequent blood meal perpetuating the transmission cycle. In Kenya, filariasis is endemic along the coastal region (3–9) and recently reported in further inland in Busia county, which is located at the Kenyan-Ugandan border (10). The main vectors of LF in Kenya are *An. gambiae* s.l, *An. funestus* and *Cx. quinquefasciatus* with varying transmission intensities which is attributed to diverse ecological and environmental conditions (6,11–14).

In 2002, the WHO launched the Global Programme to Eliminate Lymphatic Filariasis (GPELF) with the ambitious target of eliminating LF by 2020 through mass drug administration (MDA) (15). Co-administration of albendazole (400mg) and diethylcarbamazine citrate (DEC) (6 mg/kg) was recommended by the WHO for all eligible individuals in filariasis endemic areas to reduce transmission and disease morbidity. New treatment guidelines recommend a triple therapy regimen consisting of diethylcarbamazine, albendazole and ivermectin in countries without onchocerciasis (16). MDA has been tremendously successful leading to a 74% decline in LF globally (15). Kenya initiated LF elimination efforts in 2002 through annual MDA campaigns using DEC and albendazole. MDA begun in Kilifi district; a known LF foci followed by scale-up campaigns in Kwale and Malindi districts in 2003. Then to Tana river, Taita-Taveta and finally Mombasa.

The success of this strategy relies on robust surveillance and monitoring of parasite infection. Tracking data on local populations of filariasis transmitting vectors provide an opportunity for monitoring disease transmission dynamics. Monitoring of MDA performance is mainly achieved through microscopic examination of microfilariae in night time blood and detection of circulating filarial antigens. Although microscopic examinations provides the most reliable estimates, night time sampling is a major challenge and also infections may be missed in presence of unmated adult worms (17) .While monitoring of CFA can provide information about prevalence of *W. bancrofti* infection and antibody testing can provide a sensitive indicator of exposure levels, antibody testing cannot distinguish previous from current infectious leading to an over estimation of the true burden of infection. Molecular xenomontoring (MX) that relies on polymerase chain reaction (PCR) has been suggested by WHO as an important non-invasive surveillance tool to complement human surveys (17,18). MX provides a platform for monitoring infection in known vectors and also provides an opportunity to incriminate new vectors involved in the transmission of *W. bancrofti* (19). The present paper reports LF surveillance in adult mosquitoes collected on the Kenyan coast.

## Methods

### Study area

The study was conducted in the counties of Kilifi, Kwale, and Taita-Taveta along the Kenyan coast (Figure. 1). These three counties experience a moderately hot (21-31°C) and moist climate (>1000mm precipitation per year) and have a combined population of approximately 2.4 million people. Climatic changes observed in recent years include delays in the onset of rains, reduction in volume or drying up of wells and rivers, and increase in temperatures (20–22). Despite shared climatic conditions, the mosquito composition and abundance are heterogeneous with a notable decline in *An. gambiae* s.s population (23). A total of 16 sites were selected for vector sampling (Figure. 1). In Kilifi county, sampling was conducted in two former administrative units, Kilifi and Malindi district because MDA activities had previously been carried out extensively in the two regions.

**Figure 1.**
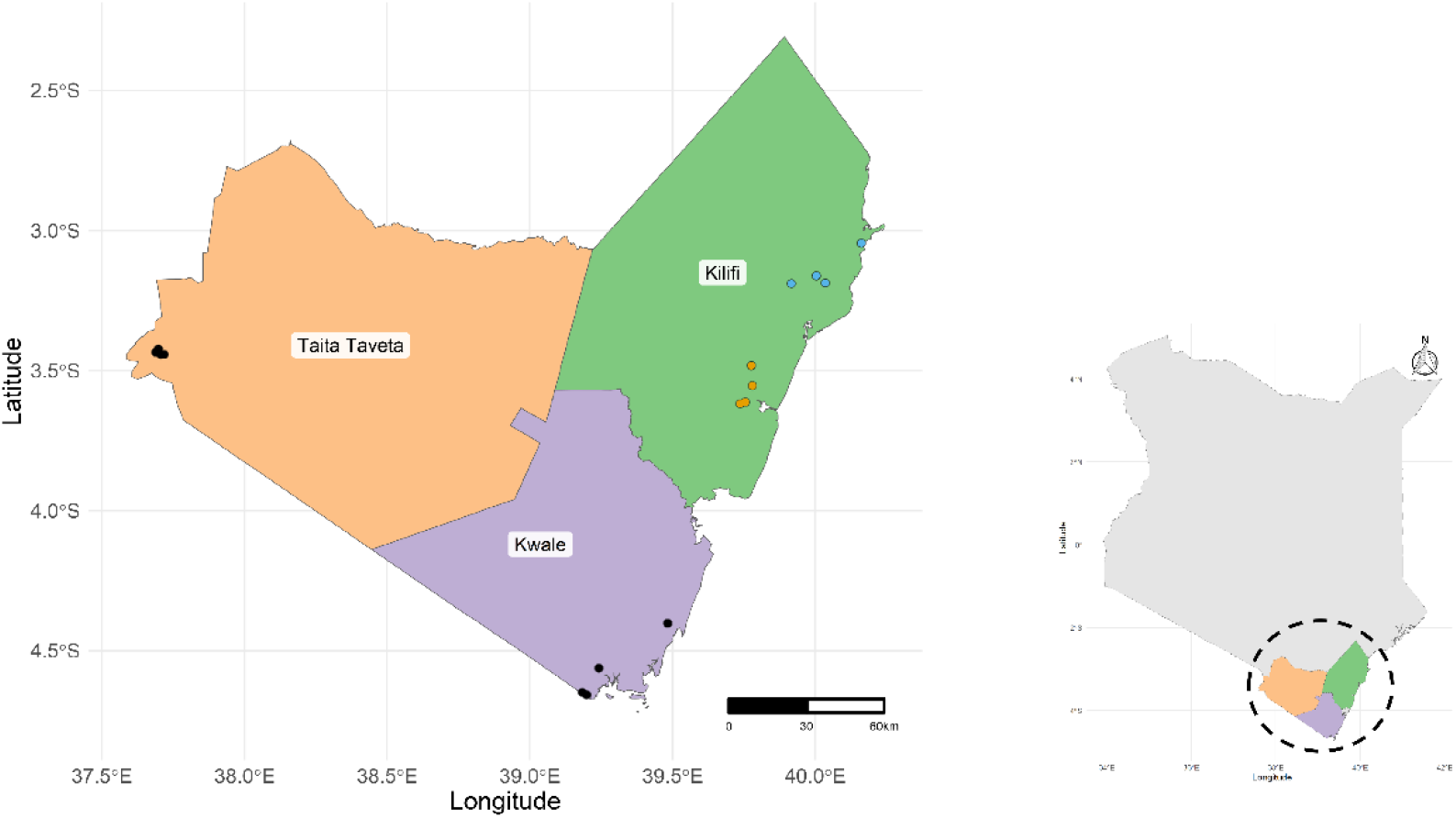
A map showing mosquito sampling sites. Sampling in Kilif county was split into two district Kilifi (gamboge) and Malindi (malibu).

### Mosquito collection

Mosquitoes collection was done both indoors and outdoors in 10 households in the 16 sites using Centers for Disease Control and Prevention light traps (CDC-LT). Sampling was carried out during the dry season (January, February and March) and at the end of the wet season (July) in 2022. The traps were set at dusk (1800hrs) and collected at dawn (0600hrs) on the next day. Geo-reference coordinates were collected using the eTrex® 10 (Garmin, Kansas, United States of America). Indoor traps were set in houses where at least one member of the household person spent the night, while the outdoor traps were placed next to livestock sheds or within a distance no more than 5 metres from the house with the indoor trap. The collected mosquitoes were identified morphologically (24) and sorted by sex and physiological state and then counted. All the Anopheline mosquitoes were preserved individually in micro centrifuge tubes containing a desiccant (silica pellets) and transported to the Kenya Medical Research Institute (KEMRI) - Wellcome Trust Research laboratory for further analysis. A small proportion of the culicine mosquitoes were collected, and the rest were discarded.

### Mosquito dissection

Using sterile scalpels and forceps, the adult female Anopheline mosquitoes were dissected into 2 parts: head and thorax and abdomen and stored at -80°C.

### DNA extraction

Genomic deoxyribonucleic acid (gDNA) was extracted from the mosquito head and thorax as described before with minor modifications (25). Briefly, sterile tungsten beads were transferred into the 1.5ml microcentrifuge tubes and topped up with 100 µl of 10% chelex and lysed using a tissue lyser at 30 hertz for 1 minute (min). The beads were removed from the tubes and the lysate incubated at 100°C for 10 min in a Thermomixer (Eppendorf, Hamburg, Germany). The solution was then centrifuged at 10,000×*g* for 2 min, supernatant transferred to a new microcentrifuge tube and stored at −80 °C.

### Molecular identification of *Anopheles gambiae* and *Anopheles funestus* sibling species

Diagnostic polymerase chain reaction (PCR) for *An. gambiae* sibling species was done using primers targeting the intergenic spacer (IGS) region of the ribosomal DNA (26) whereas for *An. funestus*, PCR primers targeting the internal transcribed spacer region 2 (ITS2) were used (27). Each individual PCR reaction consisted of 4µL 5X Green GoTaq® Flexi Buffer, 2.4µL Magnesium chloride, 4µL nuclease free water, 0.5µL Deoxynucleoside triphosphates (dNTPs), 0.1µL GoTaq® G2 Flexi DNA Polymerase, 1µL of each primer and 4µL of the mosquito DNA template. Thermocycling conditions consisted of an enzyme activation step at 95°C for 5 min, followed by 35 cycles of denaturation at 95°C for 15 seconds (secs), annealing at 55°C for 20 secs, extension at 72°C for 30 seconds (sec) and a final elongation step at 72°C for 10 mins. PCR amplicons were resolved on a 1.5% agarose gel stained using RedSafe dye and visualized using the ChemiDoc Imaging System (Bio-Rad) to resolve the different species.

### Detection of *Wuchereria bancrofti*

*Wuchereria bancrofti* was detected using the method described by Zhong et al. (28) with minor modifications. The PCR primers target the genus specific, multicopy (∼300 copies) Ssp I repeat DNA family. The PCR reaction consisted of 4µL 5X Green GoTaq® Flexi Buffer, 2.4µL Magnesium chloride, 7µL nuclease free water, 0.5µL Deoxynucleoside triphosphates (dNTPs), 0.1µL GoTaq® G2 Flexi DNA Polymerase, 1µL of each primer and 4µL of the mosquito DNA. The cycling conditions consisted of an initial enzyme activation step at 95°C for 5 min, followed by 35 cycles of denaturation at 95°C for 15 secs, annealing at 55°C for 20 secs, extension at 72°C for 30 secs and a final elongation step at 72°C for 10 mins. Amplicons were resolved on a 1.5% agarose gel stained with RedSafe dye and visualized on the ChemiDoc Imaging System (Bio-Rad). Samples with a band size of 188 base pairs were identified as positive.

### Statistical analysis

Data was entered and cleaned in a Microsoft excel file. Statistical analysis and data visualization were conducted using R software, version 4.2.1 (29). Infection proportions in the mosquito vectors were determined by dividing the number of *W. bancrofti* positive mosquitoes by the total number of mosquitoes analyzed per county in Kwale and Taita-Taveta and district in Kilifi county (Kilifi and Malindi). In order to assess the impact of various socio-demographic and household attributes on LF positivity, we employed a multilevel logistic regression model using Generalized Estimating Equations (GEE) assuming a binomial distribution. The GEE approach was chosen to account for the correlated nature of repeated observations within the regions. The model was fitted using the ‘geeglm’ function with LF positivity as the binary outcome variable. The risk factor variables included season, site of mosquito collection, mosquito species, presence or absence of eaves, livestock, poultry and bed nets, type of material used in roofs and walls and number of occupants. These factors have previously been shown to indluence the transmission of vector-borne diseases(30–33). We specified a logistic link function and selected an independence correlation structure. This was done to model within region correlations considering the the binary outcome of LF (positive or negative) and the clustered nature of the data. The results are reported as odds ratios (OR) along with 95% confidence intervals (CI) to quantify the association between the risk factor and LF positivity.

## Results

### Vector composition and abundance

A total of 18,121 mosquitoes were collected from 16 villages (Table 1). They belonged to the *Culex* (n = 11,414, 63%), Anopheles 37% (n = 6,707, 37%) genera (Table 1). The mosquitoes were predominantly caught outdoors (Figure. 2). Morphological, identification revealed that Anopheles mosquitoes consisted of *An. funestus* s.l (n = 3045, 45.4%), *An. gambiae* s.l (n = 2,873, 42.8%), *An. coustanii* (n = 662, 9.9%), *An. pharoensis* (n = 75, 1.1%), *An. maculpalpis* (n = 27, 0.4%), *An. pretoriensis* (n = 23, 0.3%) and *An. moucheti* (n = 2, 0.03%).

**Table 1.**
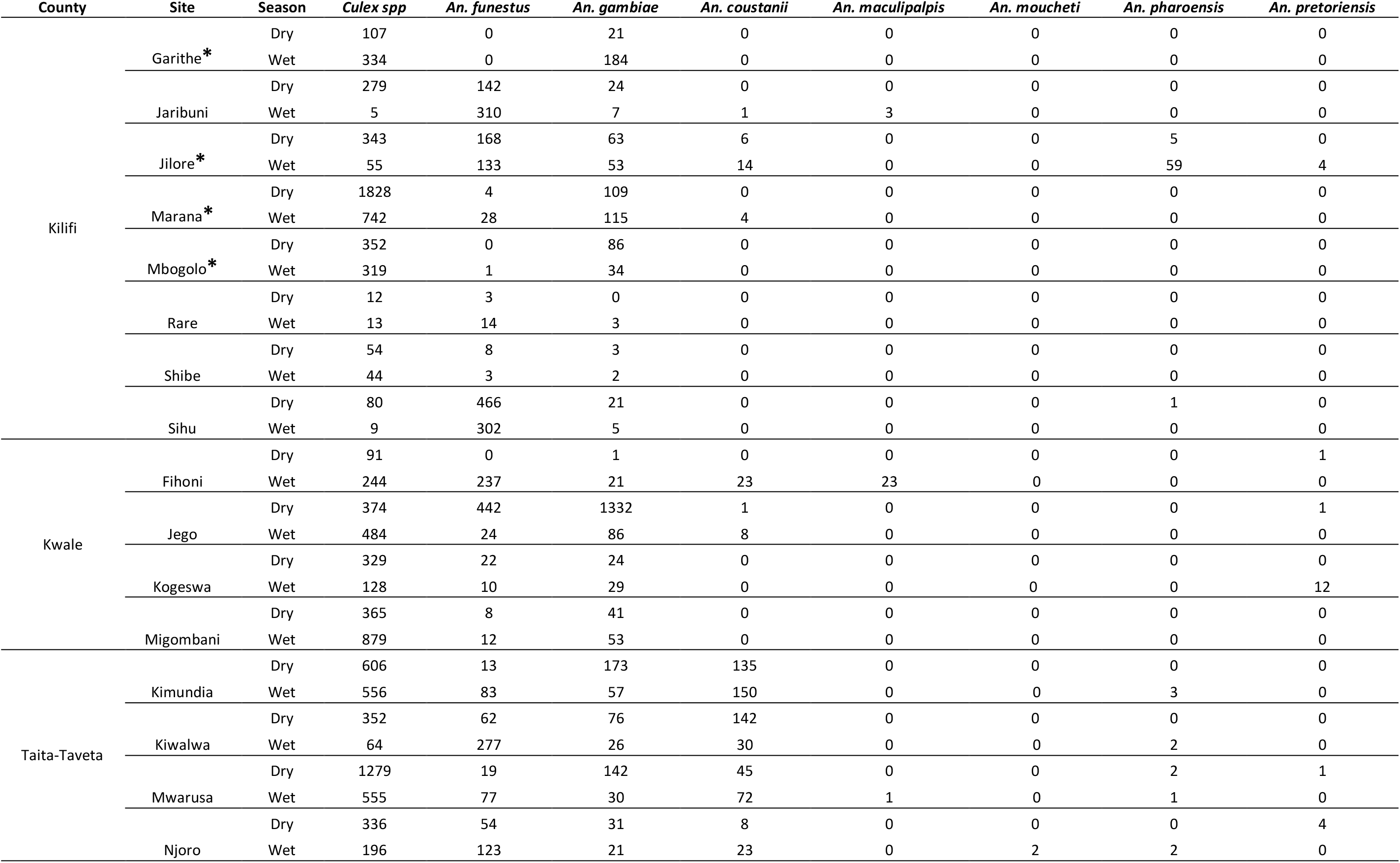
The number of morphologically identified mosquitoes collected from the 16 villages along the Kenyan coast.

**Figure 2.**
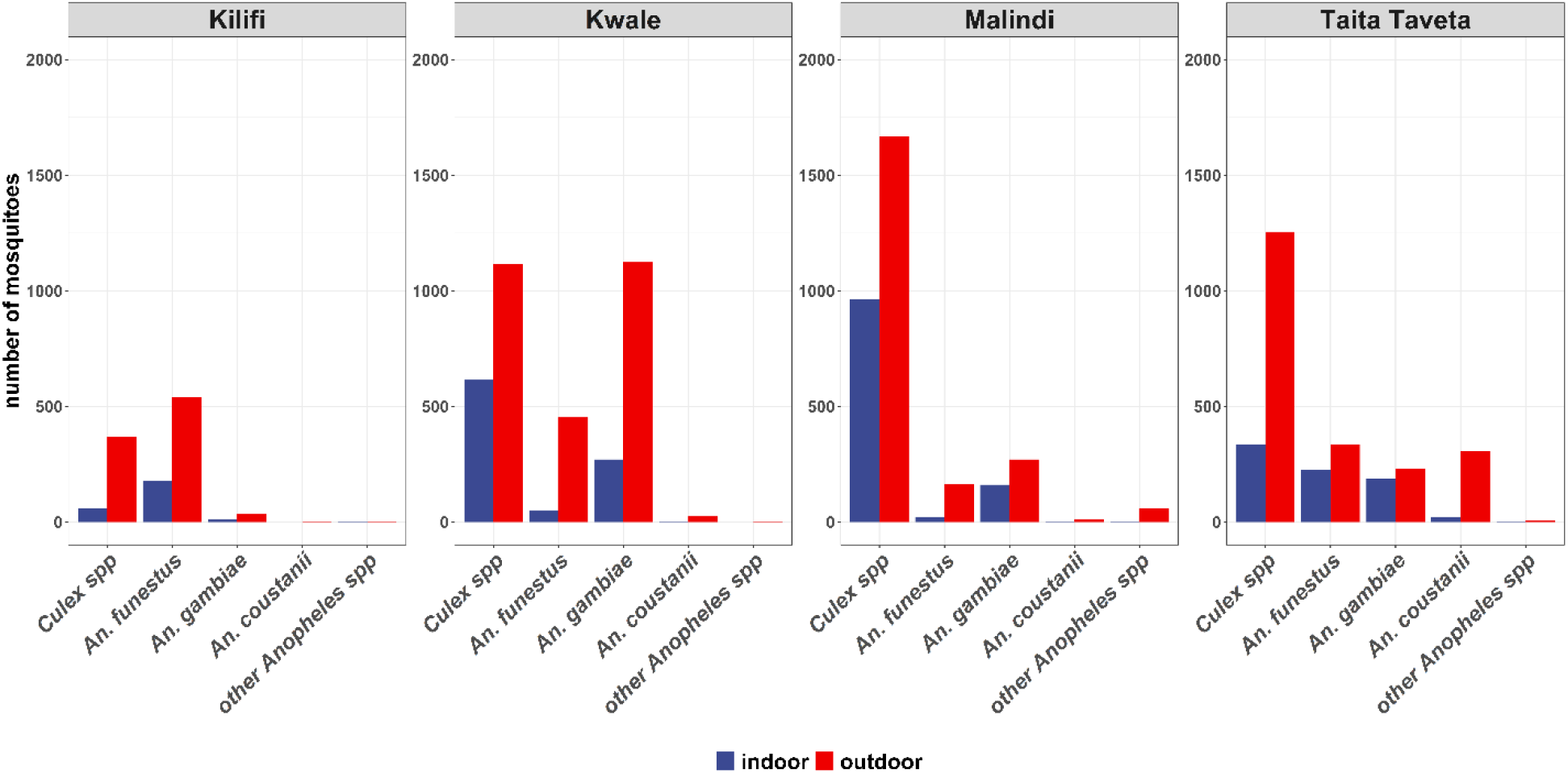
Proportion of mosquito collection indoors and outdoors.

### Bancroftian filariasis infection rates

Infection rates in the four regions were highest in Kilifi (35.4%; 95% CI 28-43.3%, n=57/161), followed by Kwale (11.7%; 95% CI 8.1-16.3%, n = 31/264), Malindi (8.3%; 95% CI 5.2-12.6%, n = 20/240) and Taita-Taveta (5.3%; 95% CI 3.4-8.0%, n = 22/412) (Table 2). In Kilifi, the highest proportions of *W. bancrofti* infected mosquitoes were *An. funestus* s.l (42.1%, n = 51) followed by *An. gambiae* s.l (15.4%, n = 6). A similar trend was observed for Malindi, Kwale and Taita Taveta. At the vector species level, parasite infection rates varied led by *An. rivulorum* (8.7%, n = 94), followed by *An. funestus s*.*s* (1.1%, n = 12), *An. arabiensis* (0.9%, n = 10) and *An. merus* (0.5%, n = 5) (Table 3). Notably, *An. rivulorum* is the dominant vector of LF in Kilifi, Kwale and Malindi, whereas in Taita-Taveta it is *An. funestus* s.s (n = 11) (Figure. 3).

**Table 2.**
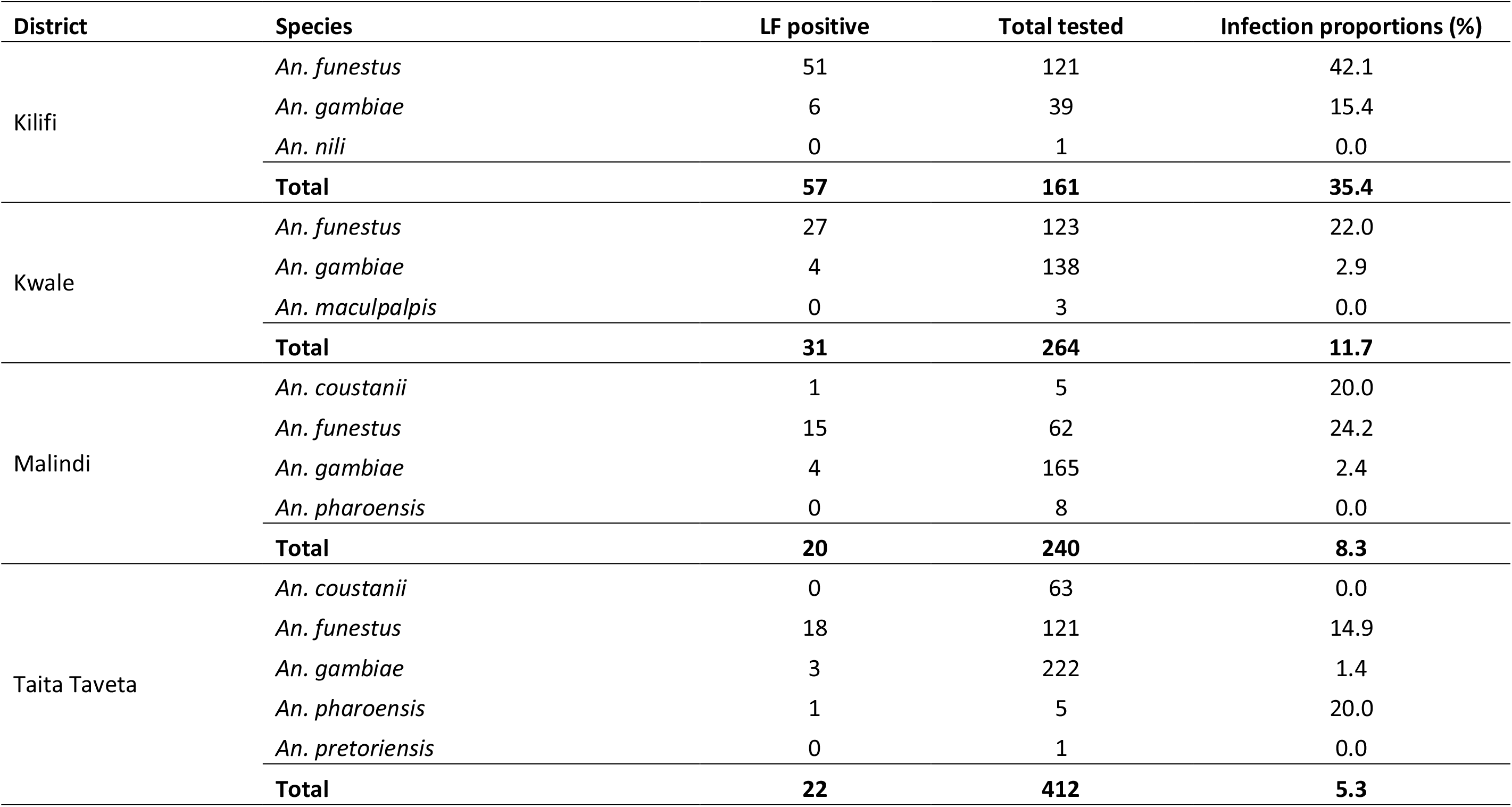
The *Wuchereria. bancrofti* prevalence rates in the three coastal counties. However, it’s important to note that Kilifi County is divided into Kilifi and Malindi districts.

| District | Species | LF positive | Total tested | Infection proportions (%) |
| --- | --- | --- | --- | --- |
| Kilifi | *An. funestus* | 51 | 121 | 42.1 |
|  | *An. gambiae* | 6 | 39 | 15.4 |
|  | *An. nili* | 0 | 1 | 0.0 |
|  | **Total** | **57** | **161** | **35.4** |
| Kwale | *An. funestus* | 27 | 123 | 22.0 |
|  | *An. gambiae* | 4 | 138 | 2.9 |
|  | *An. maculpalpis* | 0 | 3 | 0.0 |
|  | **Total** | **31** | **264** | **11.7** |
| Malindi | *An. coustanii* | 1 | 5 | 20.0 |
|  | *An. funestus* | 15 | 62 | 24.2 |
|  | *An. gambiae* | 4 | 165 | 2.4 |
|  | *An. pharoensis* | 0 | 8 | 0.0 |
|  | **Total** | **20** | **240** | **8.3** |
| Taita Taveta | *An. coustanii* | 0 | 63 | 0.0 |
|  | *An. funestus* | 18 | 121 | 14.9 |
|  | *An. gambiae* | 3 | 222 | 1.4 |
|  | *An. pharoensis* | 1 | 5 | 20.0 |
|  | *An. pretoriensis* | 0 | 1 | 0.0 |
|  | **Total** | **22** | **412** | **5.3** |

**Table 3.** *Wuchereria bancrofti* infection rates in the mosquito vectors found along Kenya coastal area.

| Species | *W. bancrofti* infection rates by PCR |  |  |  |
| --- | --- | --- | --- | --- |
|  | Positive (n) | Negative (n) | Species infection rate (%) | Overall infection rate (%) |
| *An. rivulorum*<sup>2</sup> | 94 | 193 | 32.8 | 8.7 |
| *An. funestus s.s.*<sup>2</sup> | 12 | 67 | 15.2 | 1.1 |
| *An. arabiensis*<sup>2</sup> | 10 | 359 | 2.7 | 0.9 |
| *An. merus*<sup>2</sup> | 5 | 91 | 5.2 | 0.5 |
| *An. funestus* not detected by PCR<sup>2</sup> | 3 | 18 | 14.3 | 0.3 |
| *An. lesoni*<sup>2</sup> | 2 | 23 | 8.0 | 0.2 |
| *An. gambiae* not detected by PCR<sup>2</sup> | 1 | 10 | 9.1 | 0.1 |
| *An. pharoensis*<sup>1</sup> | 1 | 12 | 7.7 | 0.1 |
| *An. coustani*<sup>1</sup> | 1 | 67 | 1.5 | 0.1 |
| *An. quadriannulatus*<sup>2</sup> | 1 | 85 | 1.2 | 0.1 |
| *An. parensis*<sup>2</sup> | 0 | 12 | 0.0 | 0.0 |
| *An. vaneedeni*<sup>2</sup> | 0 | 3 | 0.0 | 0.0 |
| *An. gambiae s.s.*<sup>2</sup> | 0 | 2 | 0.0 | 0.0 |
| *An. maculpalpis*<sup>1</sup> | 0 | 3 | 0.0 | 0.0 |
| *An. nili*<sup>3</sup> | 0 | 1 | 0.0 | 0.0 |
| *An. pretoriensis*<sup>1</sup> | 0 | 1 | 0.0 | 0.0 |
| **Total** | **130** | **947** |  | **12.1** |

**Figure 3.**
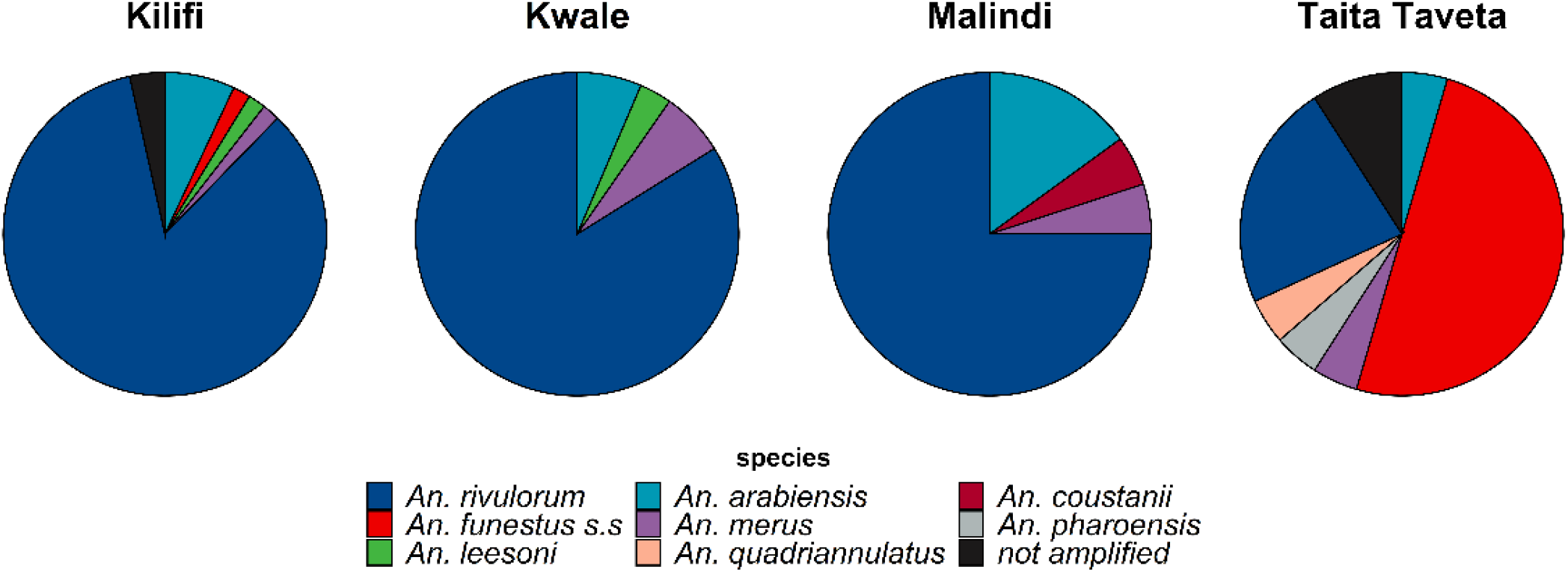
Distribution of Anopheles vectors infected with *W. bancrofti* in the Kenya Coastal region.

### Factors associated with to Bancroftian filariasis transmission

The risk of *W. bancrofti* infection was significantly higher in *An. funestus* complex (OR = 18.0; 95% CI 1.80-180, p = 0.014) compared to *An. coustanii* (Figure. 4). Additionally, higher risks were observed in outdoor resting mosquitoes (OR = 1.72; 95% CI 1.06-2.78, p = 0.027) and in homesteads that owned livestock (OR = 2.05; 95% CI 1.11-3.73, p = 0.021). Natural house construction materials used on the roof (OR = 1.49; 95% CI 0.82-2.72; p = 0.2) and wall (OR = 4.29; 95% CI 0.48-38.5; p = 0.2) were associated with increased the odds of LF transmission. Factors associated with reduced odds of infection included access to bed nets (OR=0.39; 95% CI 0.12-1.32, p = 0.13), poultry ownership (OR=0.52; 95% CI 0.30-0.89, p = 0.017) and households with fewer than two occupants (OR=0.66; 95% CI 0.41-1.08, p = 0.10).

**Figure 4.**
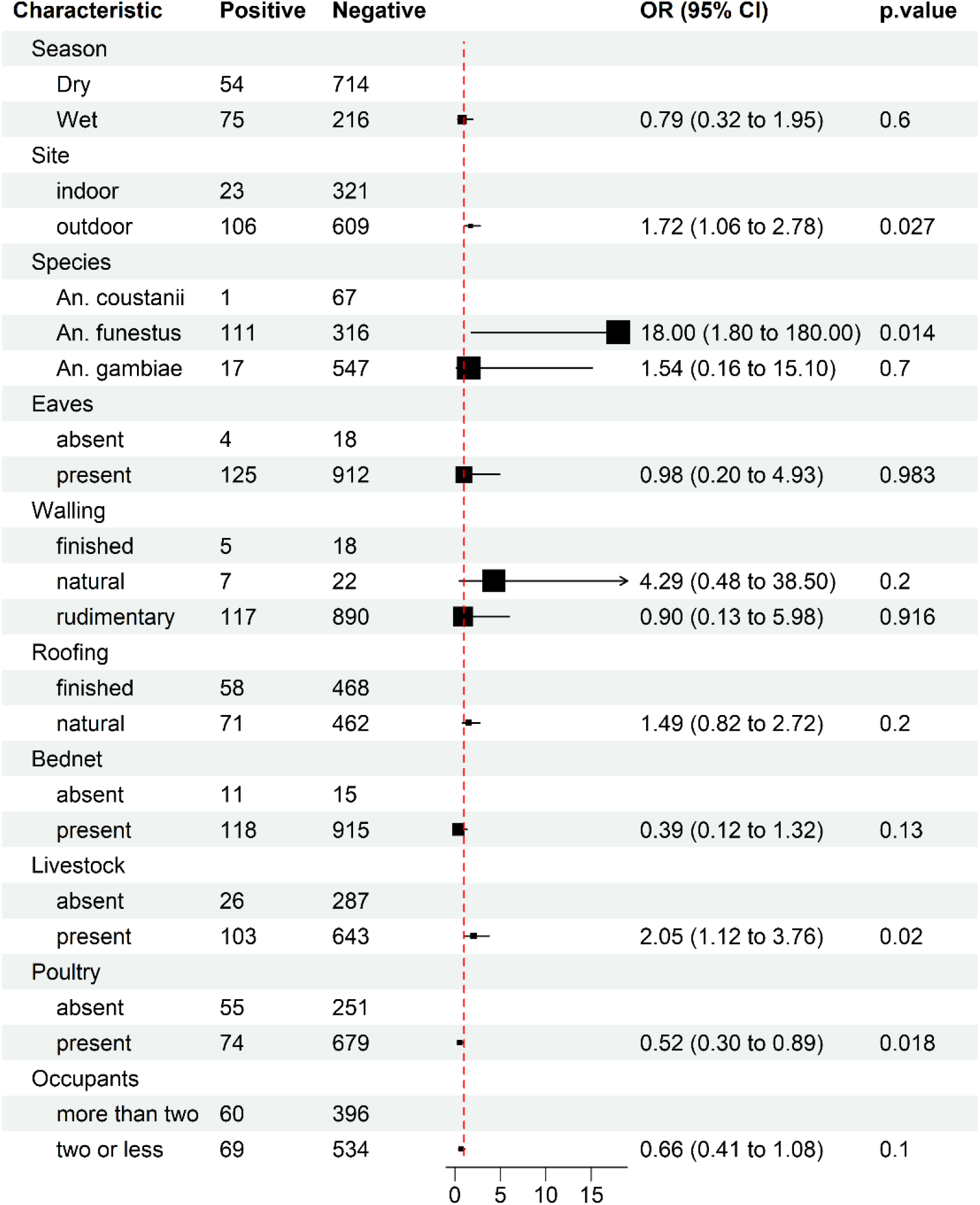
A multilevel logistic regression models based on generalized estimating equations on the various factors that may be associated with Lymphatic filariasis transmission. The outcome was either lymphatic filariasis positive or negative.

## Discussion

We investigated the vectorial systems for LF in rural coastal Kenya and factors associated with the risk of diseases transmission in the region. Efforts to eliminate LF through MDA (albendazole and diethylcarbamazine citrate) as recommended by the WHO, begun in the coastal region over two decades ago. It was carried out in subsequent years and was briefly interrupted by Covid-19. Various survey studies have been carried out in the years between, reporting CFA prevalence rates ranging from 0.3% to 6.3 % in Kwale, Kilifi and Lamu county. However, no LF cases were reported in Taita-Taveta county (7). Mosquito surveys in Malindi in 2012 showed very low prevalence where only one out of 1055 pools of mosquitoes was positive for LF (34). Using MX, we can demonstrate there’s active transmission of LF in Kilifi, Kwale and Taita-Taveta counties warranting further MDA campaigns.

Malaria and LF are co-endemic in the Kenyan coast and are transmitted by similar vectors (12,30). Over the last two decades, campaigns to control malaria, relying on long lasting insecticide bed nets (LLINs) and indoor residual sprays (IRS) have been associated with decreased malaria incidence by limiting indoor biting and resting of anthrophilic vectors, shifting vectors to more outdoor transmission (23). This study reveals a higher prevalence of LF in outdoor-resting mosquitoes, while LLIN ownership was associated with reduced risk of disease. This shows how malaria interventions may alter LF transmission dynamics by offering additional benefits that complement MDA (23,35).

*An. funestus* s.l and *An. gambiae* s.l are both involved in the transmission of LF in the study area with the former playing a much more significant role. In the *An. funestus* complex, *An. rivulorum* is the dominant vector of LF in counties adjacent to the Indian ocean whereas *An. funestus* s.s dominates further inland. In the *An. gambiae* complex, *An. arabiensis, An. merus, An. quadriannulatus* were positive for LF with no regional preference. Other mosquito species, such as *An. coustanii* and *An. pharoensis*, were indicative of LF infection, although we had very few positive samples to draw substantive conclusions about these vectors. Although high densities of *Cx. quinquefasciatus* were observed, our work did not feature this vector since its contribution to LF transmission in rural areas is still not well understood and it plays a major role in LF transmission in urban areas (36,37).

Houses made up of thatched roofs and mud walls had increased likelihood of LF transmission, this is consistent with reports in Andhra Pradesh, India (38). Such structures may provide favorable resting sites for mosquitoes. House improvements, such as screening using relatively affordable material such as papyrus mats ceilings, have been shown to reduce entry of *An. funestus* and *An. gambiae* into houses by nearly 80% in western Kenya (39). Therefore, similar modifications may be adopted to limit LF transmission in coastal Kenya.

The presence of livestock in an homestead was strongly associated with LF transmission suggesting that domestic animals play a critical role in sustaining LF vectors (35). This has been observed for Aedes albopictus that disappeared with the elimination of rats, their preferred vertebrate host (40). Therefore, incorporating vector control tools that limit access to livestock can lead to vector suppression and possible elimination of LF vectors. Presently, there are a number of studies evaluating an array of such tools including endectocides such as Ivermectin for the control of exophagic and zoophilic mosquitoes (41–43).

Interestingly, the presence of poultry within homes was associated with a lower risk of LF infections. It has previously been demonstrated that *An. arabiensis* despite feeding opportunistically on animals avoids chicken regardless of its abundance. This has been attributed to feathers acting as physical obstacles for mosquito feeding, chicken prey behavior towards mosquitoes (44), host-choice evolution driven by variation in the physical and chemical properties in the host blood or a combination of these factors (45,46). Chicken also contain volatiles (isobutyl butanoate, naphthalene, hexadecane and *trans*-limonene oxide) that repel *An. arabiensis* (44) and therefore are very beneficial addition to the peridomestic space.

## Conclusion

In this study we identified *An. funestus* s.l sibling species *An. rivulorum* and *An. funestus* s.s. as the dominant vectors of lymphatic filariasis along the Kenyan coast. We also show that a higher proportion of transmission is likely to take place outdoors necessitating the implementation of vector control strategies that target exophilic mosquitoes such as zooprophylaxis and larval source management. We also show the importance of MX in the surveillance of LF, where it’s non-invasive and has the potential for incriminating new vectors of LF.

## Data Availability

Most of the dataset used for analysis is available in the manuscript. We withheld the geo-data which may predispose individual homesteads to a high risk of identifiability. However, they are under the custodianship of the KEMRI-Wellcome Trust Data Governance Committee and are accessible upon request addressed to that committee.

## Acknowledgement

We thank the community for allowing us to conduct the research in their homes and for providing us with information. Many thanks to the technical and field staff: Festus Yaa, Gabriel Nzai, and Julius Tineja, who helped with mosquito sample collection in the field.

## Author contribution

MR designed and supervised the work. BB, KG, and JK carried out the laboratory analysis. BB, MR, LB, and AK conducted the statistical analysis. AK, CK, CM, MM, JM, and MR provided insights into the data analysis and critically reviewed the draft manuscript. All the authors read and approved the manuscript.

## Consent for publication

All the authors have reviewed and approved the publication of this paper. This paper has been published with the permission of the Director of the Kenya Medical Research Institute (KEMRI).

## Ethics declaration

The study was approved by the KEMRI Scientific and Ethics Review Unit (SERU) with the protocol number: KEMRI/SERU/CGMR-C/024/3148. Verbal informed consent was obtained from the household heads before metadata and mosquito collection.

